# Investigating the genetic relationship between vitamin B12 deficiency and Parkinson’s disease

**DOI:** 10.1101/2025.01.01.25319858

**Authors:** Raphael Dering, Margarita Onvumere, Lang Liu, Philippe Huot, Ziv Gan-Or, Konstantin Senkevich

**Author notes:** These authors equally contributed to the study (Raphael Dering, Margarita Onvumere). **Corresponding author:** Konstantin Senkevich Department of Neurology and Neurosurgery McGill University 1033 Pine Avenue, West, Ludmer Pavilion, room 309 Montreal, QC, H3A 1A1, Canada.

## Abstract

**Introduction:** Epidemiological studies suggest that patients with Parkinson’s disease (PD) may have lower levels of vitamin B12 compared to healthy controls, and it was proposed that PD patients could benefit from vitamin B12 supplementation. Functional studies have shown that B12 could modify LRRK2 activity and may directly interact with alpha-synuclein. This study aimed to investigate the role of common and rare variants in genes related to B12 metabolism and assess the potential causal relationships between B12 levels and PD risk, age-at-onset, and motor/cognitive progression.

**Methods:** We investigated the association between common and rare variants in genes involved in vitamin B12 metabolism. Rare variants (minor allele frequency < 0.01) were analyzed using the optimal sequence kernel association test (SKAT-O) in 4,815 PD patients and 65,607 controls from two independent cohorts. We constructed pathway-specific polygenic risk scores (PRS) for genes essential to B12 metabolism and for genes identified in previous genome-wide association studies (GWAS) on B12 metabolism. Mendelian randomization and genetic correlation analyses were applied to explore the relationship between vitamin B12 levels and PD risk, age-at-onset, and disease progression.

**Results:** Our analysis showed no associations between common variants of genes crucial in B12 metabolism and PD. Pathway PRS identified nominal association between B12-related genes and PD (OR = 1.061, 95% CI: 1.004–1.121, p = 0.038), which did not survive Bonferroni correction. In the rare variants analysis, we identified a significant association between variants with high CADD scores in the *CUBN* gene (P=6.07E-05; Pfdr=0.005) in the AMP-PD cohort, driven by the benign variant p.G3114S (OR=3.3; p=3.56E-05); however, this was not validated in the meta-analysis. We did not identify a potentially causal relationship between vitamin B12 levels and the risk, age-at-onset, or progression of PD. Additionally, no genetic correlation was observed between vitamin B12 and PD risk or age-at-onset GWASs.

**Conclusion:** Overall, our analyses indicate lack of genetic link between B12 levels or metabolism and PD.

## Introduction

Multiple ongoing clinical trials are investigating the efficacy of dietary adjustments in Parkinson’s disease (PD) ^1^. Several epidemiological studies have shown that vitamin B12 levels tend to be lower in patients with PD than in healthy individuals, suggesting that vitamin B12 supplementation could be beneficial in PD ^2-4^. Functional studies have demonstrated that B12 could inhibit LRRK2 kinase activity and directly interact with alpha-synuclein ^5, 6^. However, no genetic studies have causally linked B12 level or metabolism and PD or identified genetic factors leading to potentially lower B12 in PD patients ^7^.

The human body relies entirely on external sources for vitamin B12, as it cannot synthesize the vitamin internally. However, there are several genes involved in its transport and metabolism (function detailed in Table 1). These genes regulate the absorption, distribution, and conversion of B12 into its active forms ^8, 9^. Given the emerging evidence linking low B12 levels with PD, it is important to understand whether genetic variations in B12 metabolism may contribute to the risk and progression of PD.

**Table 1.** Genes involved in B12 metabolism.

| Gene | Encoded protein | Chr:position (hg38) | Function |
| --- | --- | --- | --- |
| *AMN* | Amnion associated transmembrane protein | chr14:102,922,663-102,930,842 | Part of a complex for vitamin B12 absorption in the intestines. |
| *CUBN* | Cubilin | chr10:16,823,966-17,129,811 | Receptor that binds the B12-intrinsic factor complex for absorption. |
| *MMACHC* | Methylmalonic Aciduria and Homocystinuria, Cobalamin C type | chr1:45,500,300-45,513,382 | Processes cobalamin into its active forms for cellular use. |
| *MTR* | Methionine Synthase | chr1:236,795,292-236,903,981 | Catalyzes homocysteine to methionine conversion using methylcobalamin. |
| *MTRR* | Methionine Synthase Reductase | chr5:7,869,148-7,901,113 | Regenerates methylcobalamin for methionine synthase function. |
| *TCN2* | Transcobalamin 2 | chr22:30,607,174-30,627,271 | Transports vitamin B12 in the bloodstream to tissues. |
| *FUT2* | Fucosyltransferase 2 | chr19:48,695,971-48,705,951 | Involved in the biosynthesis of the H antigen, influencing the gut microbiota and indirectly affecting vitamin B12 absorption. |

The aim of our study was to investigate the potential role of genes involved in B12 metabolism in PD and assess whether there is a potential causal relationship between B12 levels and the risk, age-at-onset, and progression of PD. First, we analyzed common and rare variants in genes related to B12 metabolism. We then constructed pathway-specific polygenic risk scores (PRS) based on the genes essential to B12 metabolism and all nominated genes identified from the previous genome-wide association studies (GWAS) on B12 metabolism. Additionally, we employed Mendelian randomization (MR) to assess whether there is a causal relationship between B12 levels and the risk, age-at-onset, and motor and cognitive progression of PD, using GWAS of B12 level as a proxy to assess the genetic determinants of B12 levels and PD risk.

## Methods

### Study Population

To investigate whether common variants in genes involved in vitamin B12 metabolism are linked to PD, we analyzed summary statistics from the largest available PD GWAS of European ancestry ^10^. Locus zoom plots were generated using the locuszoomr R library ^11^. For pathway-specific PRS analysis, seven independent cohorts were utilized, as detailed in eTable 1.

For the analysis of rare variants, we used whole-exome and whole-genome sequencing data from two independent cohorts, including 4,929 PD cases and 68,029 controls. Whole-genome sequencing data was obtained from the Accelerating Medicines Partnership – PD (AMP-PD) initiative https://amp-pd.org/; detailed in the Acknowledgments), while whole-exome sequencing data was sourced from the UK Biobank (UKBB) through the Neurohub platform https://www.mcgill.ca/hbhl/.; eTable 2).

For MR, single nucleotide polymorphisms (SNPs) derived from the GWAS on vitamin B12 serum levels were used as exposure ^12^. The largest European PD risk GWAS (excluding 23andMe, Inc. data) and a PD age-at-onset GWAS were used as outcomes ^10, 13^.

We also investigated the potential causal association between B12 and PD progression, analyzing continuous traits such as Mini-Mental State Examination (MMSE) scores, Part III Unified PD Rating Scale (UPDRS3) scores and Hoehn and Yahr Scale (HY) scores, along with binomial outcome of cognitive impairment ^14^. For the progression GWASs, sample sizes were calculated as the mean values across all SNPs in the summary statistics. All individuals in the included GWASs were of European ancestry (eTable 3). This study was approved by the Institutional Review Board of McGill University.

### Pathway-Specific Polygenic Risk Score

To investigate potential genetic association of genes involved in vitamin B12 metabolism with PD, we calculated two pathway-specific PRSs. The first PRS was based on all loci significantly associated with B12 metabolism (eTable 4) from various GWASs extracted from the GWAS Catalog ^15^. The second PRS focused solely on essential genes known to play critical roles in B12 absorption, transport, and activation (*AMN, CUBN, MMACHC, MTR, MTRR, TCN2*, and *FUT2*) were chosen because they mediate distinct steps in the conversion of B12 into its active forms in the stomach, intestine, and bloodstream ^9^ (see Table 1 for details).

We calculated PRS in participants of European ancestry from seven cohorts (eTable 1), excluding first- and second-degree relatives, as previously described ^16^. In brief, we performed a sex discrepancy check by comparing recorded biological sex with genetically inferred. For PRS calculation, common SNPs with a minor allele frequency (MAF) > 0.01 and p-value < 0.05 were included, and we performed linkage disequilibrium (LD) clumping to exclude variants with r² > 0.1 within 250kb. Covariates such as age-at-onset for PD cases, age at enrollment for controls, sex, and the top 10 principal components were included in the regression models to control for confounding factors. A permutation test with 10,000 repetitions was conducted to generate empirical p-values for the genetic pathways of interest.

### Rare variants analysis

To explore the role of rare variants in genes involved in vitamin B12 metabolism (Table 1), we utilized genetic data from whole-exome (UKBB) and whole-genome (AMP-PD) sequencing datasets. For both cohorts, genetic data were aligned to the human reference genome build hg38. We included only participants of European ancestry and excluded first- and second-degree relatives. Quality control for the whole-genome sequencing data was performed following established protocols (detailed in ^17^). Briefly, we included samples with a minimum average coverage of 25x and excluded those with more than 5% missing genotypes. For whole-exome sequencing data from the UK Biobank, we applied the Genome Analysis Toolkit (GATK, v3.8) for quality control. We included samples with minimum depth of coverage of 10x and a genotype quality (GQ) score threshold of 20.

We conducted rare variant association analysis (minor allele frequency <0.01), using the optimized sequence kernel association test (SKAT-O) ^18^. Variants were grouped into categories: all rare variants, non-synonymous variants, loss-of-function variants (stop-gain, frameshift, canonical splice-site), and variants with a Combined Annotation Dependent Depletion (CADD) score ≥ 20, predicted to be highly deleterious. Meta-analysis of the two cohorts was performed using the metaSKAT R package ^19^.

### Mendelian Randomization and Genetic Correlation

For the construction of genetic instruments in MR analysis, we extracted SNPs with GWAS-level of associations (p-value < 5.0E-08) from the exposure GWAS on B12 level ^12^. Selected SNPs were then clumped using standard parameters (clumping window of 10,000 kb, r^2^ cutoff 0.001) to exclude variants in linkage disequilibrium. To estimate the strength of genetic instruments, we calculated R^2^, which reflects the proportion of variability explained by genetic variants, and F-statistics ^20^. We then calculated power using the online Mendelian randomization power calculator to detect an equivalent effect, represented by an odds ratio (OR) of 1.2 ^21^.

To perform MR, we used the Two Sample MR R package ^22^. We performed Steiger filtering to exclude SNPs that explain more variance in the outcome than in the exposure ^23^. We then used the inverse variance weighted (IVW) method which combines results from individual Wald ratios together ^24, 25^. We applied MR Egger to identify net directional pleiotropy and obtain a more accurate estimate of the true causal effect. Heterogeneity was assessed using Cochran’s Q test. In addition, we performed an MR-PRESSO test to identify pleiotropic outlier SNPs and adjust the pooled estimate if necessary ^26^. We then performed genetic correlation analysis between PD risk and age-at-onset and B12 level GWASs, using LD Score Regression (LDSC) with standard settings ^27^.

## Results

### Genetic analysis did not identify a role for common or rare variants in genes involved in B12 metabolism in PD risk

Since the human body cannot synthesize vitamin B12, no genes directly encode for its production. Instead, we focused on key genes involved in B12 metabolism to determine whether dysfunction in these genes may contribute to risk of PD (**Table 1**). We analyzed common variants using data from the largest available PD risk GWAS ^10^ and did not find any variants reaching GWAS level of significance **(eFigure 1)**.

**Figure 1.**
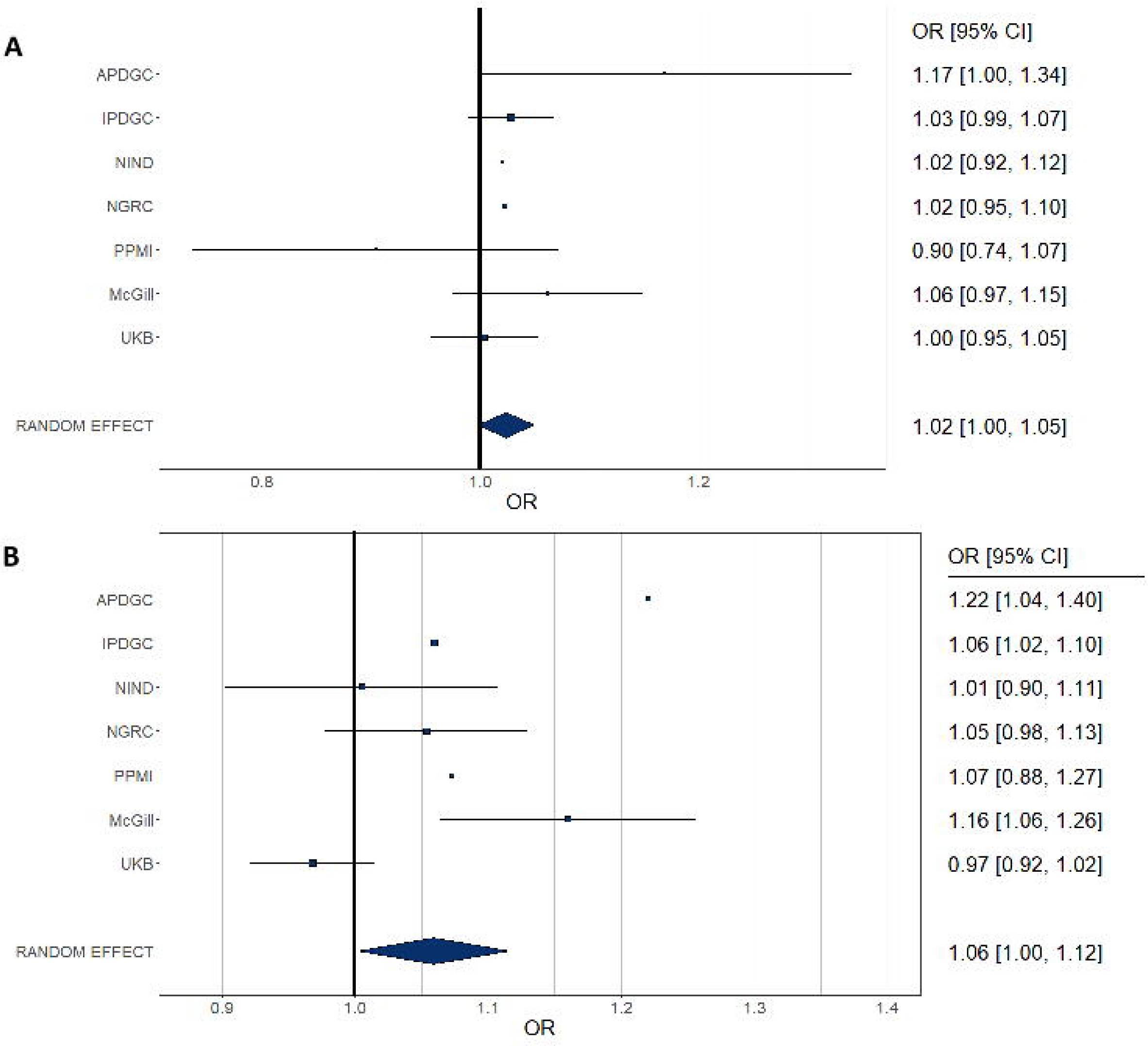
Two Pathway-Specific PRS Analyses of B12 Metabolism Genes. Panel A shows the pathway-specific PRS for 7 genes crucial in B12 metabolism. Panel B displays the pathway PRS for all GWAS-significant genes linked to B12 metabolism. Panel A shows the pathway-specific PRS for 7 genes crucial in B12 metabolism. Panel B displays the pathway PRS for all GWAS-significant genes linked to B12 metabolism. OR – Odds Ratio, CI – Confidence Interval, PPMI – Parkinson’s Progression Markers Initiative, APDGC – Autopsy-Confirmed Parkinson Disease GWAS Consortium, IPDGC – International Parkinson Disease Genomics Consortium, NINDS – National Institute of Neurological Disorders and Stroke Repository Parkinson’s Disease Collection, NGRC – NeuroGenetics Research Consortium, UKBB – UK Biobank.

We conducted two pathway PRS analyses to further evaluate the association between B12 metabolism genes and PD. The first analysis, which focused on seven key B12 metabolism genes was not significant OR=1.025 (95% CI: 0.999–1.052, p = 0.061), while the second, which included all genes reaching GWAS significance for B12 levels, showed nominal significance (OR = 1.061, 95% CI: 1.004–1.121, p = 0.038). Although both analyses suggested potential associations, these results should be interpreted with caution, as no signal remained significant after correction for multiple testing (p < 0.025; **Figure 2; eTable 5**).

We then analyzed rare variants using SKAT-O in two independent cohorts (**eTable 6**). We identified a significant association between rare variants with high CADD scores in the *CUBN* gene (P=6.07E-05; Pfdr=0.005) in the AMP-PD cohort but not in the meta-analysis after correction for multiple comparisons (P=0.011; Pfdr=0.292) with PD risk. Notably, the rare variant p.G3114S in *CUBN* was primarily responsible for this association in the AMP-PD cohort (OR=3.3; p=3.56E-05); however, this variant was not associated with PD in the UKBB cohort without proxy cases (OR=1.12; p=0.22). ClinVar classifies p.G3114S as a benign variant, with a GnomAD MAF of 0.009 in the European population. Given the lack of replication and its relatively high frequency in gnomAD, it is likely that the significance observed in the AMP-PD cohort occurred by chance. Additionally, we observed an association between high CADD score variants in *TCN2* (P=0.006; Pfdr=0.244) in the UKBB cohort, but not in the AMP-PD cohort or meta-analysis. No other associations were significant.

### Mendelian randomization and genetic correlation did not reveal causal or genetic link between B12 level and PD

The instruments had sufficient power, as indicated by F-statistics > 10. The analysis demonstrated sufficient statistical power (80%) for PD risk and age-at-onset GWASs; however, the parameters related to PD progression had less robust power, falling below the 80% threshold. No potentially causal association between B12 metabolism and PD risk, age-at-onset or progression was observed **(Table 2)**. To test for potential violations of MR assumptions, we performed sensitivity analyses **(eTable 7)**. No significant heterogeneity was evident. The results of the Egger intercept test suggest no strong evidence for horizontal pleiotropy. The MR-PRESSO test did not detect any potential pleiotropic outlier SNPs.

**Table 2.** Results of the MR analyses between B12 deficiency and PD risk and progression.

| Outcome GWAS | N SNPs | Power | Inverse variance weighted |  |
| --- | --- | --- | --- | --- |
|  |  |  | p | OR (95% CI) |
| PD | 5 | 100% | 0.892 | 1.02 (0.81-1.27) |
| PD Age at onset | 2 | 96.7% | 0.620 | 0.54 (0.05-6.27) |
| CogI | 3 | 21.5% | 0.181 | 0.33 (0.06-1.68) |
| MDS-UPDRS part III | 3 | 18.3% | 0.331 | 0.79 (0.49-1.27) |
| MMSE | 3 | 17.6% | 0.987 | 0.99 (0.43-2.3) |
| HY | 3 | 14.4% | 0.090 | 0.87 (0.73-1.02) |

R<sup>2</sup> – proportion of variance in exposure variable explained by SNPs, p – p-value, OR – Odds Ratio, CI – Confidence Interval, PD – Parkinson’s disease, CogI – Cognitive Impairment, MDS-UPDRS part III – Unified Parkinson’s Disease Rating Scale scores, MMSE – Mini-Mental State Examination scores, HY – Hoehn and Yahr Scale scores.

We did not find genetic correlation between PD risk, PD age-at-onset and B12 level GWASs (rg=-0.05, p=0.69 and rg=0.12, p=0.62, respectively; **eTable 8)**.

## Discussion

In this study, we comprehensively investigated the genetic role of B12 metabolism and levels in PD. We found no robust evidence to suggest that common or rare variants related to B12 metabolism are associated with PD risk. We also found no potential causal link between B12 levels and PD age-at-onset or risk of PD, similar to previous MR analysis ^7^ or PD progression.

Epidemiological studies suggest that B12 levels tend to be lower in PD patients as compared to healthy controls ^2-4^. It is possible that confounding factors, particularly non-motor symptoms of PD such as gastrointestinal dysfunction, weight loss with disease progression and decreased oral food intake could contribute to variations in B12 levels and may obscure any underlying associations. For example, constipation, which affects the majority of PD patients ^28^, along with delayed gastric emptying ^29^ and decreased oral intake in the later stages of the disease, could interfere with B12 levels. Additionally, alterations in gut microbiota in PD ^30^ may influence prevalence of bacteria responsible for B12 synthesis ^31, 32^.

While our analyses did not identify a genetic link, it is important to consider certain limitations. First, our analyses were restricted to individuals of European ancestry, which limits the generalizability of our findings to other populations. Second, in the MR analysis, GWAS data on PD progression have been underpowered, reducing our ability to detect associations. We were unable to access GWAS summary statistics for LRRK2-related PD, limiting our ability to assess the role of B12 in this genetic form. Thus, further studies in diverse populations and with larger datasets are needed to confirm our results.

In summary, our findings suggest that B12 levels or metabolism may not play a prominent genetic role in PD development or progression, yet addressing low B12 levels in PD patients is important to prevent B12 deficiency-related symptoms. Non-genetic factors, such as gastrointestinal dysfunction and microbiota alterations, may contribute to observed differences in B12 levels in PD patients. Future research should explore these environmental and physiological factors more closely to better understand their impact on B12 metabolism in the context of PD.

## Acknowledgments

We would like to thank the participants in the different cohorts for contributing to this study. Data used in the preparation of this article were obtained from the AMP PD Knowledge Platform. For up-to-date information on the study, visit https://www.amp-pd.org. AMP PD – a public-private partnership – is managed by the FNIH and funded by Celgene, GSK, the Michael J. Fox Foundation for Parkinson’s Research, the National Institute of Neurological Disorders and Stroke, Pfizer, Sanofi, and Verily. Genetic data used in preparation of this article were obtained from the Fox Investigation for New Discovery of Biomarkers (BioFIND), the Harvard Biomarker Study (HBS), the Parkinson’s Progression Markers Initiative (PPMI), the Parkinson’s Disease Biomarkers Program (PDBP), the International LBD Genomics Consortium (iLBDGC), and the STEADY-PD III Investigators. BioFIND is sponsored by The Michael J. Fox Foundation for Parkinson’s Research (MJFF) with support from the National Institute for Neurological Disorders and Stroke (NINDS). The BioFIND Investigators have not participated in reviewing the data analysis or content of the manuscript. For up-to-date information on the study, visit michaeljfox.org/news/biofind. The HBS is a collaboration of HBS investigators [full list of HBS investigators found at https://www.bwhparkinsoncenter.org/biobank/ and funded through philanthropy and NIH and Non-NIH funding sources. The HBS Investigators have not participated in reviewing the data analysis or content of the manuscript. PPMI – a public-private partnership – is funded by the Michael J. Fox Foundation for Parkinson’s Research and funding partners, including [list the full names of all of the PPMI funding partners found at www.ppmi-info.org/fundingpartners]. The PPMI Investigators have not participated in reviewing the data analysis or content of the manuscript. For up-to-date information on the study, visit www.ppmi-info.org. The PDBP consortium is supported by the NINDS at the National Institutes of Health. A full list of PDBP investigators can be found at https://pdbp.ninds.nih.gov/policy. The PDBP investigators have not participated in reviewing the data analysis or content of the manuscript. Genome Sequencing in Lewy Body Dementia and Neurologically Healthy Controls: A Resource for the Research Community.” was generated by the iLBDGC, under the co-directorship by Dr. Bryan J. Traynor and Dr. Sonja W. Scholz from the Intramural Research Program of the U.S. National Institutes of Health. The iLBDGC Investigators have not participated in reviewing the data analysis or content of the manuscript. For a complete list of contributors, please see: ^32^. STEADY PD III is a 36 month, Phase 3, parallel group, placebo controlled study of the efficacy of isradipine 10 mg daily in 336 participants with early Parkinson’s Disease that was funded by the NINDS and supported by The Michael J Fox Foundation for Parkinson’s Research and the Parkinson’s Study Group. The STEADY-PD III Investigators have not participated in reviewing the data analysis or content of the manuscript. The full list of STEADY PD III investigators can be found at: https://clinicaltrials.gov/ct2/show/NCT02168842. We would also like to thank the research participants and employees of 23andMe, Inc. for making this work possible. The full GWAS summary statistics for the 23andMe discovery data set will be made available through 23andMe to qualified researchers under an agreement with 23andMe that protects the privacy of the 23andMe participants. Please visit research.23andme.com/collaborate/ for more information and to apply to access the data. This research used the NeuroHub infrastructure and was undertaken thanks in part to funding from the Canada First Research Excellence Fund, awarded through the Healthy Brains, Healthy Lives initiative at McGill University, Calcul Québec and Compute Canada. This research has been conducted using the UK Biobank Resource under Application Number 45551. The UKBB cohort was accessed using Neurohub (https://www.mcgill.ca/hbhl/neurohub). ZGO is supported by the Fonds de recherche du Québec - Santé (FRQS) Chercheurs-boursiers award, and is a William Dawson Scholar.

## Data Availability

All supplementary materials, including additional tables and figures, are available in the Supplementary Materials section. The data used in the preparation of this article were obtained from the AMP PD Knowledge Platform (https://www.amp-pd.org) and the UKBB via Neurohub (https://www.mcgill.ca/hbhl/neurohub). The full GWAS summary statistics for the 23andMe inc., discovery data set will be made available through 23andMe to qualified researchers under an agreement with 23andMe that protects the privacy of the 23andMe participants. Please visit research.23andme.com/collaborate/ for more information and to apply to access the data. All code used in the analysis is available on our GitHub repository, which can be accessed at https://github.com/senkkon/B12-level-and-Parkinson-s-disease.

