## Supplemental files for "Investigating the genetic relationship between vitamin B12 deficiency and Parkinson’s disease"

###### **Supplementary Materials**

**eTable 1.** Cohorts included in the pathway Polygenic Risk Score (PRS) analysis of B12 metabolism genes.

| Cohort | N total | Case | Control | Male, N (%) |
| --- | --- | --- | --- | --- |
| APDGC | 923 | 621 | 302 | 0.65 |
| McGill | 3651 | 2381 | 1270 | 0.41 |
| PPMI | 581 | 417 | 164 | 0.67 |
| IPDGC | 10799 | 5229 | 5480 | 0.59 |
| NGRC | 3940 | 1972 | 1968 | 0.53 |
| NIND | 1686 | 896 | 790 | 0.51 |
| UKBB | 20550 | 3318 | 17232 | 0.49 |

APDGC - Autopsy-Confirmed Parkinson Disease GWAS Consortium, McGill - McGill University Parkinson’s Cohort, PPMI - Parkinson's Progression Markers Initiative, IPDGC - International Parkinson Disease Genomics Consortium, NGRC - NeuroGenetics Research Consortium, NINDS - National Institute of Neurological Disorders and Stroke Repository Parkinson’s Disease Collection, UKBB - UK Biobank

**eTable 2.** Study population for rare variant analysis.

| Cohort | All Parkinson's disease cases | | | Controls |  |  |
| --- | --- | --- | --- | --- | --- | --- |
|  | N | Mean age ± SD | Males, % | N | Mean age ± SD | Males, % |
| UKBB | 2,966 | 62.97 ± 5.26 | 1,093 (36.85%) | 64,936 | 56.78 ± 8.01 | 34,870 (53,7%) |
| AMP-PD | 1,963 | 64.54 (9.51) | 1,260 (64.2%) | 3,093 | 69.76 (13.02) | 1,496 (48.4%) |
| All cohorts | 4,929 | - | - | 68,029 | - | - |

UKBB - UK Biobank, AMP-PD - Accelerating Medicines Partnership - Parkinson’s Disease Initiative, SD - Standard deviation

**eTable 3.** GWAS studies included in the Mendelian randomization analysis.

| Trait | Trait category | GWAS study | N cases | N controls | Sample size |
| --- | --- | --- | --- | --- | --- |
| B12 level | Continuous | Dennis et al., 2021 | - | - | 19,415 |
| PD | Binomial | Nalls et al., 2019 | 33,674 | 449,056 | 482,730 |
| PD Age at onset | Continuous | [Blauwendraat et al., 2019](https://pubmed.ncbi.nlm.nih.gov/30957308/) | - | - | 17,996 |
| CogI | Binomial | [Iwaki et al., 2019](https://pubmed.ncbi.nlm.nih.gov/31505070/) | - | - | 1,710 |
| MDS-UPDRS part III | Continuous | [Iwaki et al., 2019](https://pubmed.ncbi.nlm.nih.gov/31505070/) | - | - | 1,398 |
| MMSE | Continuous | [Iwaki et al., 2019](https://pubmed.ncbi.nlm.nih.gov/31505070/) | - | - | 1,329 |
| HY | Continuous | [Iwaki et al., 2019](https://pubmed.ncbi.nlm.nih.gov/31505070/) | - | - | 1,005 |

For CogI, MDS-UPDRS part III, MMSE, and HY GWASs, sample sizes were calculated as mean values across all SNPs in the summary statistics, PD - Parkinson’s disease, CogI - Cognitive Impairment, MDS-UPDRS part III - Unified Parkinson’s Disease Rating Scale part III, MMSE - Mini-Mental State Examination, HY - Hoehn and Yahr Scale.

**eTable 4.** Genes reported in the association with B12 level in previous GWAS studies and used to construct pathway PRS.

| CUBN | SLC25A2 | TCN2 |
| --- | --- | --- |
| TCN1 | LINC01881 | ABCD4 |
| RASIP1 | CASC17 | MMAA |
| FUT2 | CYP3A54P | ELAPOR2 |
| PRELID2 | LINC02253 | ALG1L9P |
| APTX | CFAP299 | FUT3 |
| MS4A3 | RGS7 | TAF7 |
| CLYBL | RN7SKP181 | CALM2P1 |
| FUT6 | SRD5A3P1 | SDK1 |
| MMUT | CD320 | OOSP1 |

**eTable 5.** Pathway polygenic risk score analysis of the B12 metabolism genes

| Cohort | Set | PRS.R2 | PRS.R2.adj | Full.R2 | Null.R2 | P | Num_SNP | Empirical-P |
| --- | --- | --- | --- | --- | --- | --- | --- | --- |
| APDGC | B12_all_genes | 0.011 | 0.005 | 0.052 | 0.048 | 0.008 | 78 | 0.015 |
| IPDGC | B12_all_genes | 0.001 | 0.000 | 0.012 | 0.012 | 0.003 | 13 | 0.006 |
| McGill | B12_all_genes | 0.006 | 0.002 | 0.198 | 0.196 | 0.000 | 79 | 0.001 |
| NGRC | B12_all_genes | 0.001 | 0.000 | 0.139 | 0.139 | 0.159 | 80 | 0.293 |
| NIND | B12_all_genes | 0.000 | 0.000 | 0.060 | 0.060 | 0.920 | 78 | 0.993 |
| PPMI | B12_all_genes | 0.001 | 0.001 | 0.032 | 0.031 | 0.447 | 19 | 0.690 |
| UKB | B12_all_genes | 0.003 | 0.002 | 0.175 | 0.173 | 0.099 | 96 | 0.187 |
| Meta-analysis | Random effect model |  | OR=1.061 | 95%CI (1.004-1.121) | | **0.038** |  |  |
| Cohort | Set | PRS.R2 | PRS.R2.adj | Full.R2 | Null.R2 | P | Num_SNP | Empirical-P |
| APDGC | B12_7genes | 0.007 | 0.003 | 0.051 | 0.048 | 0.038 | 10 | 0.071 |
| IPDGC | B12_7genes | 0.000 | 0.000 | 0.012 | 0.012 | 0.157 | 8 | 0.287 |
| McGill | B12_7genes | 0.001 | 0.000 | 0.196 | 0.196 | 0.156 | 10 | 0.283 |
| NGRC | B12_7genes | 0.000 | 0.000 | 0.139 | 0.139 | 0.542 | 10 | 0.785 |
| NIND | B12_7genes | 0.000 | 0.000 | 0.060 | 0.060 | 0.706 | 10 | 0.911 |
| PPMI | B12_7genes | 0.003 | 0.001 | 0.033 | 0.031 | 0.286 | 6 | 0.490 |
| UKB | B12_7genes | 0.004 | 0.003 | 0.175 | 0.173 | 0.075 | 12 | 0.146 |
| Meta-analysis | Random effect model |  | OR=1.025 | 95%CI (0.999-1.052) | | **0.061** |  |  |

OR – Odds Ratio, CI – Confidence Interval, PPMI – Parkinson's Progression Markers Initiative, APDGC – Autopsy-Confirmed Parkinson Disease GWAS Consortium, IPDGC – International Parkinson Disease Genomics Consortium, NINDS – National Institute of Neurological Disorders and Stroke Repository Parkinson's Disease Collection, NGRC – NeuroGenetics Research Consortium, UKBB – UK Biobank.

**eTable 6.** Burden analysis of rare variants in genes involved in B12 metabolism

| **Cohorts** | SetID | P.value | N.Marker.Test | MAC | Pfdr |
| --- | --- | --- | --- | --- | --- |
| AMP_PD | CUBN_CADD | **6.07E-05** | 29 | 185 | 0.005 |
| AMP_PD | CUBN_nonsyn | 0.052 | 120 |  | 0.980 |
| AMP_PD | MMACHC_LOF | 0.088 | 3 | 10 | 0.872 |
| AMP_PD | AMN_nonsyn | 0.110 | 5 | 149 | 0.970 |
| AMP_PD | FUT2_All | 0.159 | 78 | 979.5749 | 1.000 |
| AMP_PD | MMACHC_CADD | 0.187 | 15 | 35.0002 | 1.000 |
| AMP_PD | MTRR_LOF | 0.253 | 1 | 1 | 1.000 |
| AMP_PD | MMACHC_nonsyn | 0.304 | 21 | 69.0004 | 1.000 |
| AMP_PD | FUT2_nonsyn | 0.342 | 19 | 100 | 1.000 |
| AMP_PD | AMN_LOF | 0.361 | 1 | 89 | 1.000 |
| AMP_PD | MTR_CADD | 0.391 | 7 | 19 | 1.000 |
| AMP_PD | MTR_All | 0.398 | 659 | 8663.55 | 1.000 |
| AMP_PD | MTRR_All | 0.413 | 274 | 2967.251 | 0.988 |
| AMP_PD | CUBN_All | 0.427 | 1729 | 21219.05 | 0.911 |
| AMP_PD | MTR_nonsyn | 0.479 | 20 | 169.0004 | 0.945 |
| AMP_PD | MTR_LOF | 0.495 | 2 | 2 | 0.931 |
| AMP_PD | AMN_All | 0.497 | 74 | 940.1064 | 0.913 |
| AMP_PD | CUBN_LOF | 0.507 | 5 | 58 | 0.911 |
| AMP_PD | FUT2_LOF | 0.519 | 1 | 1 | 0.892 |
| AMP_PD | TCN2_All | 0.611 | 170 | 2758.172 | 0.878 |
| AMP_PD | MTRR_CADD | 0.624 | 7 | 69 | 0.864 |
| AMP_PD | TCN2_nonsyn | 0.665 | 17 | 146 | 0.876 |
| AMP_PD | MMACHC_All | 0.697 | 92 | 1074.004 | 0.874 |
| AMP_PD | MTRR_nonsyn | 0.749 | 26 | 155 | 0.846 |
| AMP_PD | TCN2_CADD | 0.761 | 4 | 75 | 0.835 |
| UKBB | TCN2_CADD | 0.006 | 4 | 14.00034 | 0.244 |
| UKBB | MTRR_nonsyn | 0.054 | 175 | 1642.529 | 0.859 |
| UKBB | MTRR_All | 0.172 | 471 | 5972.683 | 1.000 |
| UKBB | MTRR_CADD | 0.176 | 56 | 1048.153 | 1.000 |
| UKBB | TCN2_nonsyn | 0.257 | 31 | 203.006 | 1.000 |
| UKBB | CUBN_nonsyn | 0.289 | 627 | 8448.732 | 1.000 |
| UKBB | MTR_All | 0.309 | 524 | 7020.389 | 1.000 |
| UKBB | MTRR_LOF | 0.356 | 8 | 41.00909 | 1.000 |
| UKBB | FUT2_nonsyn | 0.401 | 72 | 1142.002 | 1.000 |
| UKBB | CUBN_CADD | 0.426 | 120 | 2250.646 | 0.934 |
| UKBB | MMACHC_CADD | 0.467 | 23 | 340.0206 | 0.946 |
| UKBB | FUT2_LOF | 0.511 | 4 | 14.00002 | 0.897 |
| UKBB | MMACHC_nonsyn | 0.548 | 37 | 475.0199 | 0.921 |
| UKBB | TCN2_All | 0.552 | 196 | 958.1867 | 0.909 |
| UKBB | FUT2_CADD | 0.609 | 2 | 6 | 0.907 |
| UKBB | MTR_nonsyn | 0.609 | 172 | 2900.355 | 0.891 |
| UKBB | CUBN_LOF | 0.650 | 19 | 81.17143 | 0.870 |
| UKBB | MMACHC_All | 0.691 | 97 | 1226.036 | 0.895 |
| UKBB | MTR_LOF | 0.697 | 5 | 12.00129 | 0.887 |
| UKBB | MMACHC_LOF | 0.707 | 4 | 6.003401 | 0.859 |
| UKBB | AMN_All | 0.726 | 225 | 5951.892 | 0.869 |
| UKBB | FUT2_All | 0.744 | 134 | 1435.39 | 0.877 |
| UKBB | AMN_nonsyn | 0.745 | 50 | 2219.219 | 0.853 |
| UKBB | AMN_CADD | 0.790 | 4 | 4.000092 | 0.844 |
| UKBB | TCN2_LOF | 0.805 | 2 | 3 | 0.826 |
| UKBB | MTR_CADD | 1.000 | 91 | 663.27 | 1.000 |
| meta-analysis | CUBN_CADD | **0.011** |  |  | 0.292 |
| meta-analysis | CUBN_nonsyn | 0.073 |  |  | 0.961 |
| meta-analysis | MMACHC_LOF | 0.076 |  |  | 0.858 |
| meta-analysis | MTRR_LOF | 0.194 |  |  | 1.000 |
| meta-analysis | MTRR_nonsyn | 0.224 |  |  | 1.000 |
| meta-analysis | CUBN_All | 0.273 |  |  | 1.000 |
| meta-analysis | MTR_All | 0.279 |  |  | 1.000 |
| meta-analysis | FUT2_All | 0.334 |  |  | 1.000 |
| meta-analysis | AMN_LOF | 0.361 |  |  | 1.000 |
| meta-analysis | AMN_All | 0.406 |  |  | 1.000 |
| meta-analysis | FUT2_LOF | 0.407 |  |  | 1.000 |
| meta-analysis | AMN_nonsyn | 0.417 |  |  | 0.970 |
| meta-analysis | MTR_LOF | 0.423 |  |  | 0.955 |
| meta-analysis | MTRR_CADD | 0.428 |  |  | 0.890 |
| meta-analysis | MMACHC_CADD | 0.490 |  |  | 0.944 |
| meta-analysis | TCN2_All | 0.586 |  |  | 0.945 |
| meta-analysis | MTR_nonsyn | 0.595 |  |  | 0.940 |
| meta-analysis | CUBN_LOF | 0.605 |  |  | 0.937 |
| meta-analysis | FUT2_CADD | 0.609 |  |  | 0.925 |
| meta-analysis | TCN2_nonsyn | 0.621 |  |  | 0.876 |
| meta-analysis | TCN2_CADD | 0.640 |  |  | 0.872 |
| meta-analysis | MMACHC_nonsyn | 0.701 |  |  | 0.865 |
| meta-analysis | FUT2_nonsyn | 0.744 |  |  | 0.865 |
| meta-analysis | MMACHC_All | 0.753 |  |  | 0.837 |
| meta-analysis | MTRR_All | 0.785 |  |  | 0.849 |
| meta-analysis | AMN_CADD | 0.790 |  |  | 0.832 |
| meta-analysis | TCN2_LOF | 0.805 |  |  | 0.837 |
| meta-analysis | MTR_CADD | 0.863 |  |  | 0.874 |

**eTable 7.** Heterogeneity tests and tests for directional horizontal pleiotropy of the MR analyses between B12 deficiency and PD risk and progression.

| Trait | Heterogeneity tests | | | | | | Tests for directional horizontal pleiotropy | | | |
| --- | --- | --- | --- | --- | --- | --- | --- | --- | --- | --- |
|  | Inverse variance weighted | | | MR Egger | | | egger intercept | SE | p | MR-PRESSO Global Test P-value |
|  | Q | Q_df | Q_pval | Q | Q_df | Q_pval |  |  |  |  |
| PD | 2.91 | 4 | 0.574 | 0.59 | 3 | 0.898 | -0.07 | 0.05 | 0.226 | 0.735 |
| PD Age-at-onset | 1.54E-03 | 1 | 0.969 | NA | NA | NA | NA | NA | NA | NA |
| CogI | 0.85 | 2 | 0.654 | 0.69 | 1 | 0.406 | 0.11 | 0.28 | 0.759 | NA |
| MDS-UPDRS part III | 0.52 | 2 | 0.770 | 0.26 | 1 | 0.608 | -0.04 | 0.08 | 0.700 | 0.950 |
| MMSE | 1.87 | 2 | 0.393 | 1.02 | 1 | 0.313 | 0.12 | 0.13 | 0.529 | 0.661 |
| HY | 0.51 | 2 | 0.773 | 0.35 | 1 | 0.554 | 0.01 | 0.03 | 0.755 | 0.615 |

NA for Q test and horizontal pleiotropy tests if not enough instrumental variables; **Abbreviations**: Q = Cochran's Q test value; Q_Df = Q degrees of Freedom; Q_p = Q p-value; B12 = B12 deficiency level; PD = Parkinson’s disease; CogI = Cognitive Impairment; MDS-UPDRS part III = Part III of the Unified Parkinson's Disease Rating Scale scores; MMSE = Mini-Mental State Examination scores; HY = Hoehn and Yahr Scale scores.

**eTable 8.** Genetic Correlation Analysis between GWAS on B12 Deficiency and Parkinson's Disease and its Age at Onset Risk.

| Genetic correlation of B12 level GWAS with | Genetic Correlation (SE) | Z-score | P-value |
| --- | --- | --- | --- |
| PD risk | -0.049 (0.122) | -0.398 | 0.691 |
| PD age at onset | 0.120 (0.248) | 0.489 | 0.625 |

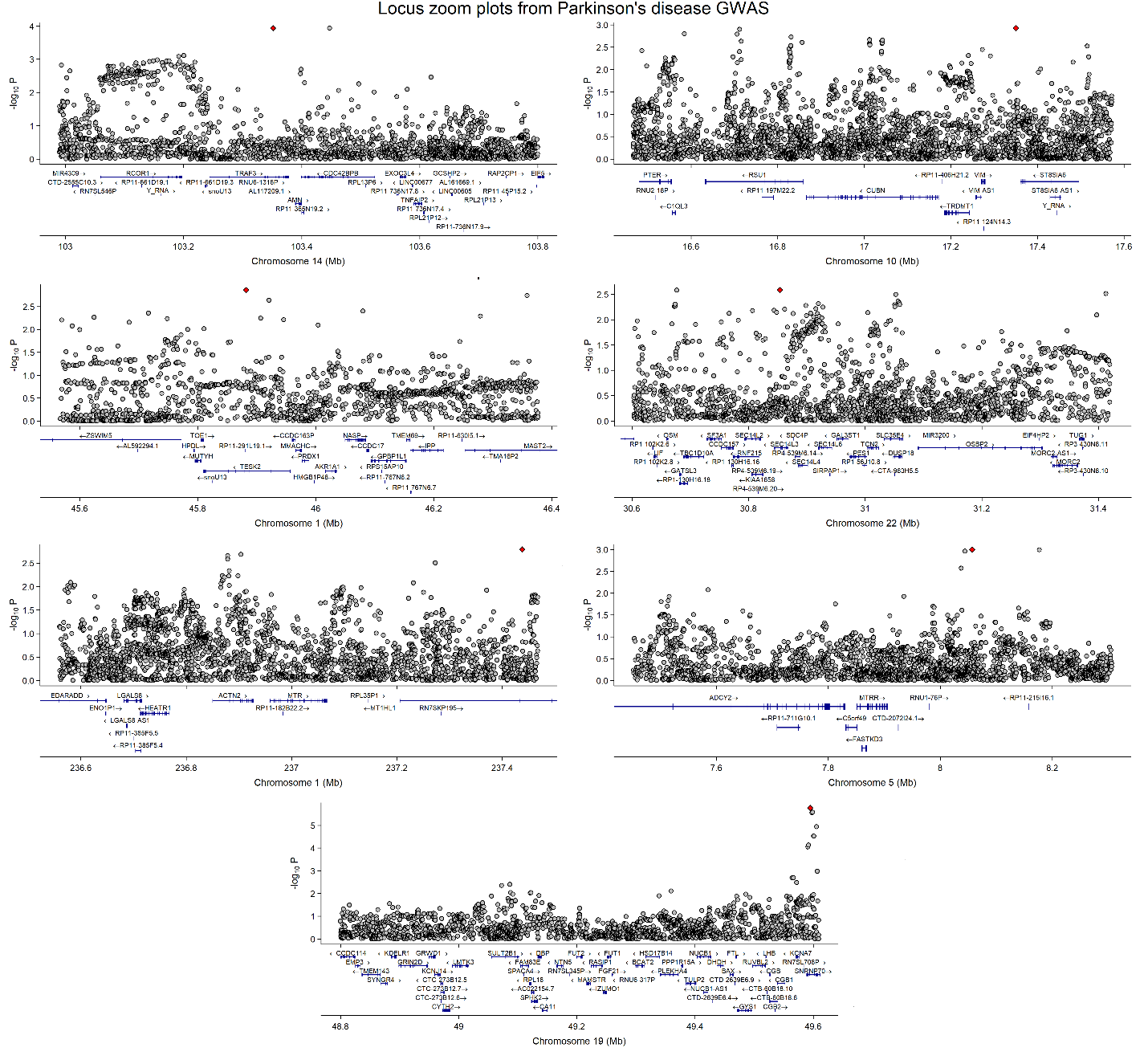

eFigure 1**. Locus zoom plot of genes related to B12 metabolism (*AMN, CUBN, MMACHC, MTR, MTRR, TCN2*, and *FUT2;* +/- 500 kb) from a Parkinson’s disease GWAS.**

The SNP with the lowest P-value for each gene or locus is highlighted with a red square. None of the SNPs in any of the loci reached genome-wide significance (p < 5 × 10⁻⁸), with all SNPs shown in grey falling below this threshold. X-axis: Chromosomal position (in megabases). Y-axis: Negative log10 of the P-values from the GWAS.
